# Echinacea purpurea for the Long-term Prevention of Viral Respiratory Tract Infections during COVID-19 Pandemic: A Randomized, Open, Controlled, Exploratory Clinical Study

**DOI:** 10.1101/2021.12.10.21267582

**Authors:** Emil Kolev, Lilyana Mircheva, Michael R. Edwards, Sebastian L. Johnston, Krassimir Kalinov, Rainer Stange, Giuseppe Gancitano, Wim Vanden Berghe, Samo Kreft

## Abstract

**Introduction:** SARS-CoV-2 vaccination is effective in preventing severe COVID-19, but efficacy in reducing viral load and transmission wanes over time. In addition, the emergence of novel SARS-CoV-2 variants increases the threat of uncontrolled dissemination and additional antiviral therapies are urgently needed for effective containment. In previous *in vitro* studies *Echinacea purpurea* demonstrated strong antiviral activity against enveloped viruses, including SARS-CoV-2. In this study, we examined the potential of *Echinacea purpurea* in preventing and treating respiratory tract infections (RTIs) and in particular, SARS-CoV-2 infections.

**Methods:** 120 healthy volunteers (m,f, 18 – 75 years) were randomly assigned to *Echinacea* prevention or control group without any intervention. After a run-in week, participants went through 3 prevention cycles of 2, 2 and 1 months with daily 2’400mg *Echinacea purpurea* extract (Echinaforce®, EF). The prevention cycles were interrupted by breaks of 1 week. Acute respiratory symptoms were treated with 4’000 mg EF for up to 10 days, and their severity assessed via a diary. Naso/oropharyngeal swabs and venous blood samples were routinely collected every month and during acute illnesses for detection and identification of respiratory viruses, including SARS-CoV-2 via RT-qPCR and serology.

**Results:** Summarized over all phases of prevention, 21 and 29 samples tested positive for any virus in the EF and control group, of which 5 and 14 samples tested SARS-CoV-2 positive (RR=0.37, Chi-square test, p=0.03). Overall, 10 and 14 symptomatic episodes occurred, of which 5 and 8 were COVID-19 (RR=0.70, Chi-square test, p>0.05). EF treatment when applied during acute episodes significantly reduced the overall virus load by at least 2.12 log_10_ or approx. 99% (t-test, p<0.05), the time to virus clearance by 8.0 days for all viruses (Wilcoxon test, p=0.02) and by 4.8 days for SARS-CoV-2 (p>0.05) in comparison to control. Finally, EF treatment significantly reduced fever days (1 day vs 11 days, Chi-square test, p=0.003) but not the overall symptom severity. There were fewer COVID-19 related hospitalizations in the EF treatment group (N=0 vs N=2).

**Discussion/Conclusion:** EF exhibited antiviral effects and reduced the risk of viral RTIs, including SARS-CoV-2. By substantially reducing virus loads in infected subjects, EF offers a supportive addition to existing mandated treatments like vaccinations. Future confirmatory studies are warranted.

**Clinical Trials registration Nr:** NCT05002179

## Introduction

Respiratory tract infections (RTI) represent the most frequent illness in western civilization [1]. Especially during winter months, a plethora of endemic viruses causes substantial pressure to individuals and the health care system [2]. While common non-influenza illnesses are a massive burden on society and economies, completely novel types of pathogen (variants of influenza or coronaviruses) pose a great threat to humanity. As such, COVID-19 presents the latest and certainly most significant coronavirus zoonosis in the last 20 years.

Initial efficacy studies on COVID-19 vaccines raised high hopes of curbing the pandemic through vaccination endeavors. Messenger RNA and vector-based vaccines showed >90% effectiveness in preventing overall infections, progression to severe illness as well as transmission of SARS-CoV-2 [3, 4] Expectedly, infection protective effects seemed to slowly reduce over time manifested by increasing breakthrough infections observed even in fully vaccinated individuals [5, 6]. The emergence of novel SARS-CoV-2 mutations, e.g. the delta variant featuring higher peak virus loads and transmissibility than previous variants presents another threat to containment by immunization [7, 8]. Most recent data from US Health Administration, relating to 2.7% of the US population found vaccine effectiveness declining from 87.9% to 48.1% from February to October 2021, with great differences between applied vaccines. Prevention of severe Covid-19 illness remained high throughout the time post vaccination and irrespective of virus mutation in contrast to overall SARS-CoV-2 infections and viral loads, both correlated with the risk of transmitting infections [9]. Additional options are urgently needed to effectively attenuate non-severe infections and naso-oropharyngeal virus concentrations in order to further contain viral dissemination [10].

Broad antiviral effects, including virucidal activity against coronaviruses (CoV) were attributed to the medicinal plant *Echinacea* [11-13]. *In vitro*, a hydroethanolic extract prepared from freshly-harvested herb and root parts of *Echinacea purpurea* (Echinaforce®, EF) inhibited infectivity of human CoV 229E, highly pathogenic MERS- and SARS-CoV, as well as the newly identified SARS-CoV-2 [11]. Two earlier prevention studies in adults and children suggested clinically relevant benefits of EF for enveloped viral pathogens including coronaviruses [14]. The same extract exhibited adaptive immuno-modulating properties *in vivo* by reducing the inflammatory cytokines TNF and IL-1β and increasing the anti-inflammatory cytokine IL-10 [15]. Immunomodulation instead of immune-stimulation can allow, if necessary, a prolonged preventive use of this extract, to exploit its potential ability to reduce viral loads.

This exploratory study aimed to determine antiviral effects of EF during the Covid-19 pandemic and found that the extract potently reduced SARS-CoV-2 infections and viral loads as part of an overall effect on viral respiratory tract infections.

## Methods

### Study Design and Participants

This randomized, parallel, open, no-treatment controlled, exploratory study was carried out in Bulgaria from 30^th^ of November 2020 (first patient first visit) to 29^th^ of May 2021 (last patient last visit) at one study centre (Diagnostics and Consultation Center Convex EOOD, Sofia). Principally healthy subjects residing in Sofia and neighboring regions were recruited from the principal investigator’s database and through referrals. Subjects provided written consent prior to their participation and assignment to either the Echinacea or control group. This study was carried out in compliance with ICH-GCP and according to the Declaration of Helsinki (2013). It was approved by the local ethical review board (Ethics Committee at Diagnostics and Consultation Center Convex Ltd, Sofia, registration nr: 116/26.10.2020) and registered on clinicaltrials.gov (identifier: NCT05002179).

The following exclusion criteria applied: age <18 years, >75 years, positive pregnancy test/no contraception, long-term intake of antimicrobials/antivirals/immune-suppressors, surgical intervention within 3 months prior to study or planned, diabetes mellitus, bronchopulmonary dysfunctions/diseases, immune system/metabolic disorders, serious health conditions, known allergies to ingredients of study medication, participation in clinical study within 30 days prior to study or planned.

After a run-in observation week, participants in the verum group went through 3 prevention cycles of 2, 2 and 1 months (Figure 1) with 3 times daily 800 mg EF extract (2’400 mg/day). We chose 1-week breaks for treatment interruption following regulatory advice, although the duration of pausing was not officially stipulated.

**Figure 1.**
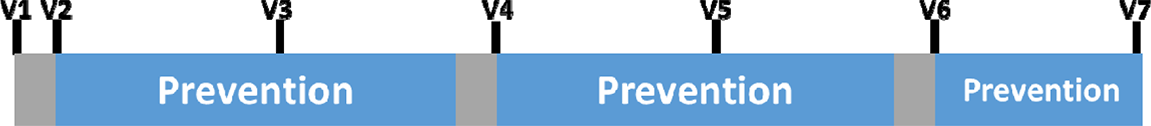
Illustration of the intervention scheme showing phases of EF prevention (blue) interrupted by phases of breaks (grey) with study visits (V1-V7) for routine virus sampling.

Acute RTI episodes were treated with five times daily 800mg EF extract (4’000 mg/day) for up to 10 days. In the control group, EF was not applied neither as prevention nor as therapy, but subjects were observed in parallel for the same period. Subjects were randomly assigned to study groups according to the randomization list (generated by SAS®/PLAN module). Beside the preventive intake of EF in the verum group, participants were allowed to continue previous treatment and therapies throughout the study and were free to use concomitant treatment during acute RTI episodes. Any concomitant treatment was recorded.

Subjects returned to the study centre on a monthly basis (visits V1-V7, Figure 1) and during acute symptomatic episodes on days 1, 2, 5 and 10 to provide naso/oropharyngeal (NP/OP) swabs and venous blood samples for virus detection and quantification. Detections, pre-existing at V1 or occurring during the run-in phase before the start of prevention at visit 2 were not taken into account for the analysis of incidence rates. The study nurse visited SARS-CoV-2 positive subjects who were confined to domestic quarantine in accordance with local law, to collect NP/OP and blood samples every 5^th^ day until they were tested SARS-CoV-2 negative. Venous blood samples (7.0 mL) were additionally drawn for analysis of serology (see below).

Subjects were equipped with a symptom diary to rate the severity of respiratory symptoms (runny nose, congested nose, sneezing, cough, shivering, malaise, fatigue, headache, myalgia, anosmia, insomnia, sore throat) upon occurrence and for up to 10 days using a Likert scale [absent=0 to severe=3] and body temperature [°C, arm pit measurement] according to Jackson (1958) [16]. Adverse events (AE) during study conduct were collected via patient diary and during study visits, classified according to preferred / lowest-level term, severity and causal relationship by the investigator. AEs were coded according to the MedDRA (version 17.1GE). Concomitant medication use was collected via patient diary and during study visits and classified according to WHO ATC, L3 Code.

### Laboratory procedures

Nasopharyngeal (NP) and oropharyngeal (OP) swabs for general RTI viral detection were collected using sterile FLOQ swabs (COPAN SA, Italy) and transferred to eNAT medium tube (COPAN SA, Italy). Sample preparation and reverse transcriptase – quantitative polymerase chain reaction (RT-qPCR) measurement were done using VIASURE RT-PCR detection kit for respiratory viruses using the Respiratory Panel IV (CerTest BIOTEC S.L., Spain). The collected samples were screened for presence of rhinoviruses, enteroviruses, adenoviruses and enveloped viruses including: influenza A (including H1N1)/B, parainfluenza 1/2/3/4, respiratory syncytial virus A/B, coronaviruses: 229E/NL63/OC43/HKU1, metapneumovirus and bocavirus.

NP and OP swabs for SARS-CoV-2 detection were collected using sterile polyurethane foam bud Σ-Transwabs (Medical Wire & Equipment (MWE), United Kingdom) with breakpoints, pooled and transferred to one tube of 1 mL Amies liquid culture medium (MWE, United Kingdom). Sample preparation and RT-qPCR measurement were done using a separate SARS-CoV-2 panel (Taqpath Covid 19 –ThermoFisher Scientific, USA). An additional serological analysis of venous blood samples was carried out for qualitative detection of SARS-CoV-2 IgG/IgM done with the Elecsys Anti SARS-CoV-2 kit (Roche Diagnostics Int., Switzerland).

All samples were stored at -80 °C until further processing at the study centre and analyzed by Bodimed diagnostic laboratories (Sofia, Bulgaria) in strict adherence to the manufacturer’s diagnostic protocols. Virus presence was detected by RT-qPCR in NP/OP swabs and serology. Cycle threshold values (Ct) were deducted from RT-qPCR measurements to estimate relative differences of virus genome copies, i.e. the virus load. SARS-CoV-2 S-, N- and ORF1ab-genes cycle threshold values (Ct) were pooled for further mathematical analysis of virus loads (see below).

### Intervention

Echinaforce® tablets (EF) used in this study contained 400mg of liquid extract (extraction solvent 65% v/v ethanol) of freshly harvested *Echinacea purpurea* (95% aerial parts and 5% root, DER = 1:11-12) and excipients. The tablets were placed into dark brown glass bottles with a screw closure and sealed. Each bottle contained 120 tablets sufficient for 20 days of prevention. Good manufacturing practice (GMP)-compliant manufacturing and batch-release was performed by A. Vogel AG (Roggwil, Switzerland). Each included subject randomized into the EF prevention group received a number of glasses sufficient for each prevention cycle. Compliance was determined based on weighing returned study product glasses upon end of prevention cycles and a tolerance of ±20% accepted for adherence to therapy.

### Sample size calculation & statistics

This study principally used descriptive biometric approaches to estimate effect sizes. However, the study was conceptualized and large enough to confirm a clinically relevant difference for a first parameter in hierarchy of pre-defined variables, i.e. incidences of viral respiratory tract infections (RTIs), with appropriate statistical power (nQuery Advisor, 2017, version 7.0, sample size and power calculation from Statsols-Statistical Solutions Ltd, IRL): A two group Chi-square test with a two-sided significance level (α = 0.05) had 80% power to detect a difference in RTI incidence rate of 0.12, with absolute rates of 0.10 in the verum group and 0.23 for control, when the sample size in each group was at least 50. In this study, we planned to recruit and observe N=120 healthy volunteers, equally randomized (Echinacea/verum group: N=60 and control group: N=60).

Relative log_10_ change in virus load after 5 and 10 days of treatment compared to baseline (day 1) during RTI episodes was calculated by approximation from the Cycle threshold values (Ct) of RT-qPCR measurements in accordance with methods described elsewhere [[17, 18]. Ct values of treatment responders falling below the detection limit of the respective RT-qPCR assay were set to the maximal number of cycles run in the respiratory qPCR panel = 45 Ct, and of the SARS-CoV-2 qPCR panel = 40 Ct. Subsequently missing Ct values due to hospitalization of severe COVID-19 (2 cases) were replaced by the last observation carried forward principle up do day 10.

Safety variables were analyzed in the safety group (SAF), which included all subjects with at least one documented intake of the study medication. Analyzes of effectiveness variables were carried out on the intention to treat group (ITT), which included all subjects with at least one evaluable effectiveness variable. Thus in this study, the ITT group was identical to the SAF group.

Continuous variables were expressed descriptively and post-hoc comparison tests carried out as indicated. Relative risk (RR) and odds ratio (OR) were adjusted for the relative subject observation time in order to take into account different observation periods in both study groups due to some participants not undertaking prevention cycle 3. Adjusted RR and OR were displayed with their 95% confidence intervals (CI). Two-sided p-values less than 0.05 were considered statistically significant. All statistical analyses were done using the SAS® system (version 9.4).

## Results

### Baseline characteristics

Overall, N=120 volunteers were included into the clinical trial in November/December 2020 and observed over a period of 23 weeks, resp. 5.5 months, as shown in Figure 1. 100% were Caucasian with a mean age of 36 years, a high proportion of smokers (36.7%) and with average body measures as shown in Table 1. 37 subjects (30.8%) had positive RT-qPCR or serology detections for SARS-CoV-2 upon inclusion (EF:20, control:17, p>0.05). Rates of smokers, overall co-morbidities and in particular hypertension in particular were slightly higher in the EF group as shown in Table 1. Otherwise, the two study groups were comparable.

**Table 1.**
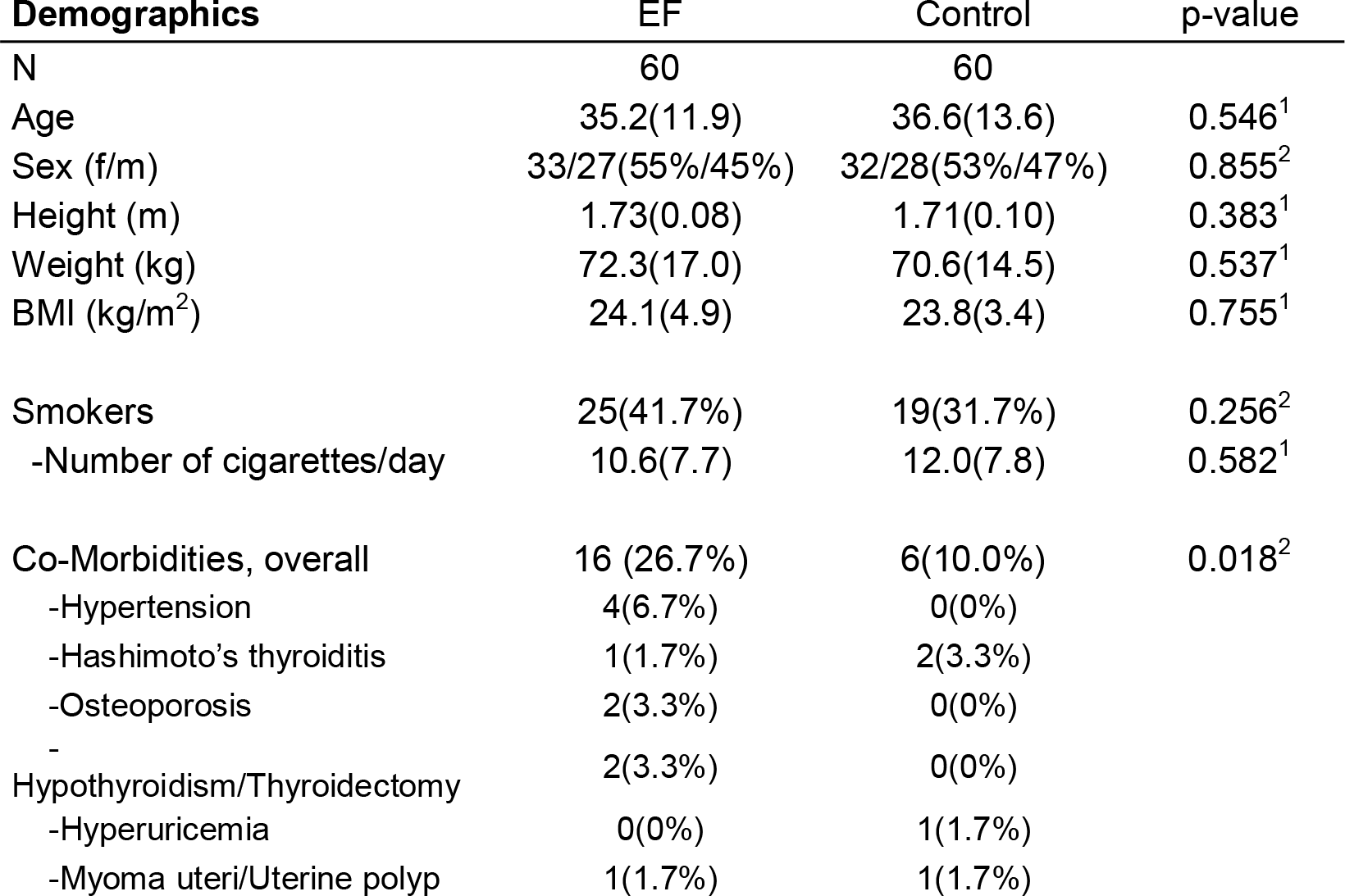
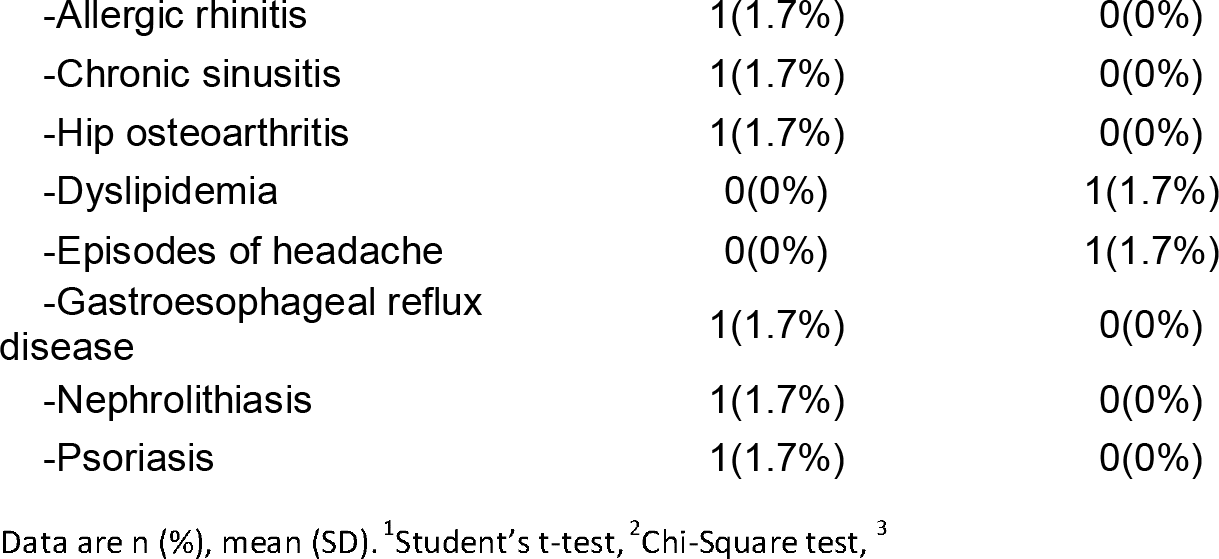
Demographics and co-morbidities.

As depicted in the (consort) flow diagram (Figure 2), N=2 subjects were screening failures due to violation of in/exclusion criteria and N=120 subjects were ultimately randomized. One participant (1.67%) of the EF group dropped out prior taking any study medication and 1 more (1.67%) during study conduct. N=58 (96.7%) in the EF group and N=60 (100%) in the control group completed the two prevention cycles (2 and 2 months). An amendment to the study allowed to voluntarily extend the initially approved 2 × 2 prevention cycles by another month of prevention in both study groups. N=49 (81.7%) and 59 (98.3%) decided to complete prevention cycle 3 (1 month). Dropouts in this study were not replaced. All subjects that decided to revoke their consent during conduct of the study provided evaluable datasets until the time point of withdrawal. The overall subject observation time in the EF and control group was 1252 and 1316 subject-weeks, respectively (ratio: 0.951).

**Figure 2.**
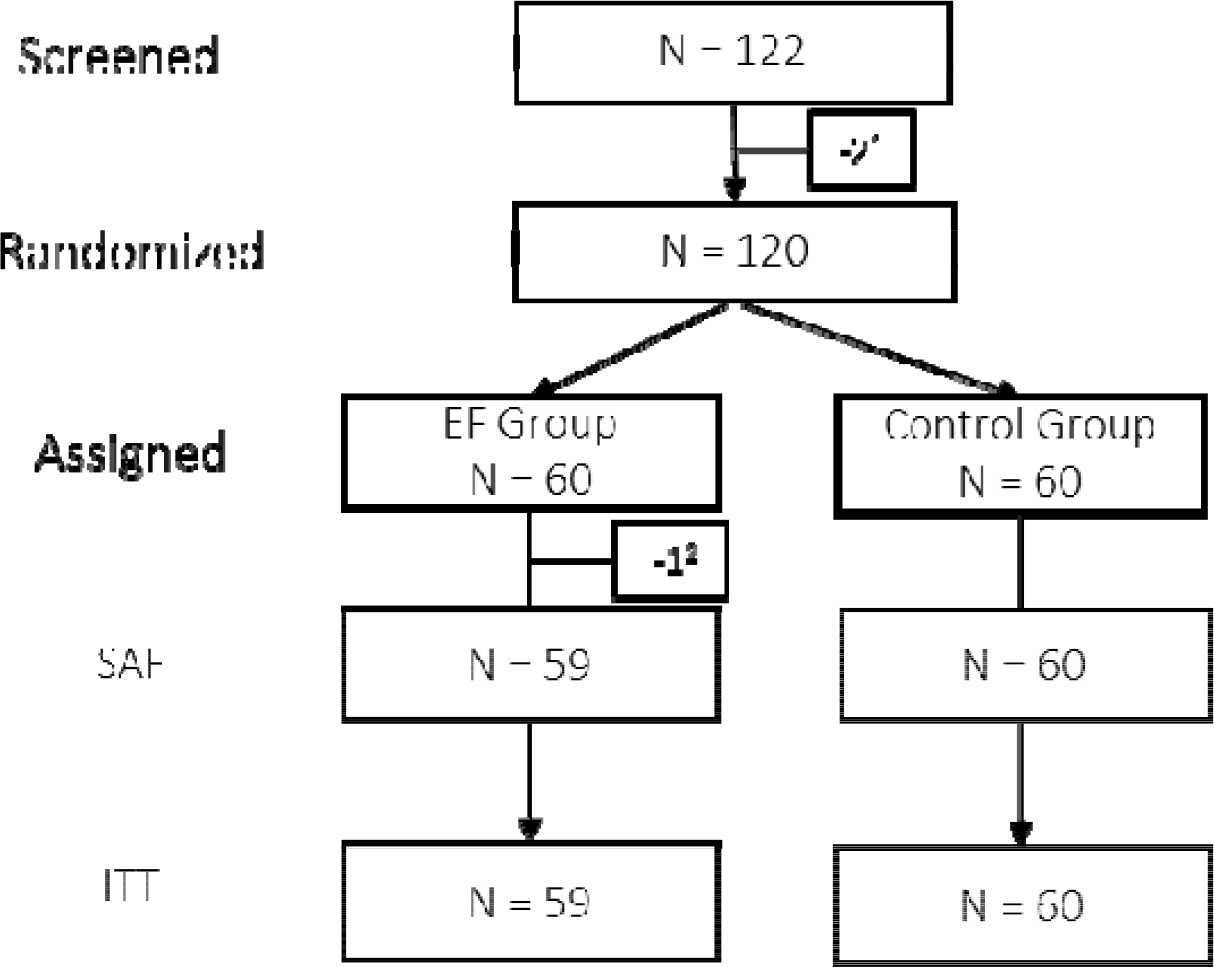
Subject disposition tree. ^1^Screening-failure, ^2^Withdrawal of consent prior intake of study medication, SAF: Safety group, ITT: Intention to treat group.

Overall, 59 (EF) and 60 subjects (control) contributed datasets evaluable for efficacy and safety variables (SAF/ITT). At study start, no subject was vaccinated against COVID-19. Three subjects (EF: 2, control: 1) received a first SARS-CoV-2 vaccine dose towards the end of the first prevention phase (prior visit 4). As few as 12 subjects (EF: 7, control: 5) received complete vaccination by the end of the study (prior visit 7). Overall, no significant differences between groups were detected and treatment compliance was 92% (95%CI: 89%/95%) in the treatment group.

### Incidence of Viral Respiratory Tract Infections and SARS-CoV-2

Table 2A shows the incidences of positive virus detections, measured by RT-qPCR and/or serology during phases of prevention with EF or the matching observation period in the control group. An overall antiviral effect is evident from the 21 (EF) and 29 (control) samples positively tested for any respiratory virus. It reveals an accentuated specificity towards enveloped viruses, with 11 (EF) and 20 (control) positive detections, which finally peaks in 5 (EF) and 14 (control) SARS-CoV-2 positive detections, respectively. The corresponding relative risk (RR.) reduced from RR = 0.748 for any respiratory virus (p=0.186), to a statistically significant RR = 0.517 for coronaviruses (p=0.046) and RR = 0.369 for SARS-CoV-2 virus infections (p=0.030) (Table 2A). EF prevention thus resulted in a virus protective effect size of 25% (relative risk reduction) for any virus, of 48% for coronaviruses and of 63% for SARS-CoV-2 virus in particular.

**Table 2.** Incidences of RTI virus detections (A) and symptomatic RTI episodes (B) during phases of prevention.

| **(A)**<br>**RTI Virus**<br>**detections** | EF | Control | OR.<sup>1</sup> | 95%CI for<br>OR | RR<sup>1</sup> | 95%CI for RR | p-value<sup>2</sup> |
| --- | --- | --- | --- | --- | --- | --- | --- |
| All RTI viruses | 21 | 29 | 0.61 | 0.29/1.26 | 0.748 | 0.49/1.16 | 0.186 |
| Enveloped<br>viruses | 11 | 20 | 0.47 | 0.20/1.09 | 0.568 | 0.30/1.09 | 0.077 |
| Coronaviruses | 10 | 20 | 0.42 | 0.18/0.995 | 0.517 | 0.27/1.02 | 0.046 |
| SARS-CoV-2 | 5 | 14 | 0.31 | 0.1/0.92 | 0.369 | 0.14/0.96 | 0.03 |
| **(B)**<br>**Symptomatic**<br>**RTI episodes** | EF | Control | OR | 95%CI for<br>OR | RR | 95%CI for<br>RR | p-value<sup>2</sup> |
| Overall<sup>3</sup> | 10 | 14 | 0.71 | 0.29/1.76 | 0.77 | 0.37/1.60 | 0.34 |
| All RTI viruses | 7 | 10 | 0.73 | 0.26/2.05 | 0.77 | 0.31/1.89 | 0.433 |
| Enveloped viruses | 6 | 9 | 0.68 | 0.23/2.05 | 0.72 | 0.28/1.91 | 0.389 |
| Coronaviruses | 6 | 9 | 0.68 | 0.23/2.05 | 0.72 | 0.28/1.91 | 0.389 |
| SARS-CoV-2 | 5 | 8 | 0.66 | 0.20/2.15 | 0.70 | 0.24/2.03 | 0.396 |

<sup>1</sup>OR: Odds ratio (OR) /risk ratio (RR) adjusted for relative subject-observation time, <sup>2</sup>Chi-square test, <sup>3</sup>incl. symptomatic episodes without any RTI virus detection.

Preventive effects of EF observed at the level of symptomatic respiratory tract infection episodes (RTI episodes) seemed to point into the same direction (Table 2B). The overall relative risk to encounter symptomatic RTI episodes was reduced by 23%, respectively 30% for episodes caused by SARS-CoV-2. Although showing highly similar possible effect sizes, the study was ultimately underpowered to show statistical significance at this level, as only every third virus infections turned into a symptomatic RTI episode.

During the two weeks of break between prevention cycles, respiratory viruses were present in 10 and 5 samples in the EF prevention- and control group, respectively (p=0.05, Chi-square test). 5 were endemic pathogens (CoV-NL63 [3], parainfluenza [1], rhinovirus [1]) and as few as 6 (EF prevention) and 4 (control) SARS-CoV-2 infections occurred, all of which remained asymptomatic. These detections were not included in the primary analysis, which focused on incidences during the treatment periods with Echinaforce.

### Virus Concentration in Oro-/Nasopharyngeal Samples

During symptomatic RTI episodes, oro-/nasopharyngeal sampling was intensified to determine virus loads and time to virus clearance in EF treatment and control groups. While initial virus loads (Ct values) on day 1 of RTI episodes were comparable (Table S1A), we found evidence for significantly more efficient reduction of virus load under EF treatment relative to baseline (Tables 2 & S1B). After 5 and 10 days, EF treatment reduced overall virus concentration significantly in comparison to day 1, while under control it remained unchanged until day 5. For both time points, the log_10_ΔCt reduction was with -2.12 (95%CI: -0.90/-3.34, t-test, p=0.0018) and -2.82 (95%CI: -1.04/-4.59, t-test, p=0.0327) higher under EF treatment (Table 2) in comparison to control. This corresponded to a significant >99% reduction in relative virus concentration. Highly comparable and equally significant results were obtained for SARS-CoV-2 virus loads with observed log_10_ΔCt reductions of -2.18 (day 5, 95%CI: - 0.77/-3.58, t-test, p=0.0054) and -2.21 (day 10, 95%CI: -0.12/-4.29, t-test, p=0.0399) in comparison to control.

**Table 2.** Log change in virus load during EF treated (Echinaforce) vs. untreated (control) symptomatic RTI episodes.

|  | All Viral RTI Episodes |  | SARS-CoV-2 episodes |  |
| --- | --- | --- | --- | --- |
|  | day 5 | day10 | day 5 | day10 |
| <u>EF</u> |  |  |  |  |
| n | 11 | 11 | 8 | 8 |
| Mean log<sub>10</sub>ΔCt | -2.19(1.33) | -4.73(1.91) | -2.14(1.28) | -3.92(1.47) |
| Median | -2.38 | -4.59 | -2.22 | -4.25 |
| 95%CI for Mean | -3.08/-1.30 | -6.01/-3.45 | -3.21/-1.07 | -5.15/-2.68 |

| Control |  |  |  |  |
| --- | --- | --- | --- | --- |
| n | 9 | 9 | 7 | 7 |
| Mean $\log_{10}\Delta Ct$ | -0.07(1.26) | -1.91(1.85) | 0.03(1.24) | -1.71(2.08) |
| Median | 0 | -2.2 | 0 | -1.9 |
| 95%CI for Mean | -1.04/0.90 | -3.33/-0.49 | -1.11/1.18 | -3.63/0.21 |
| p-value<sup>1</sup> | 0.0018 | 0.0327 | 0.0054 | 0.0399 |

Data are mean(SD) change in logarithmized $\Delta Ct$ values by day 5 and 10 of treatment relative to baseline on day 1 ( $\log_{10}\Delta Ct$ )
<sup>1</sup>Welch's t-test using Satterthwaite modification comparing EF treatment vs control.

All SARS-CoV-2 infections were followed-up every 5 days after day 10 until naso-/oropharyngeal samples tested negative. Compared to control, EF treatment significantly shortened the average time to virus clearance (qPCR-negative) by 8.02 days for all viruses (95%CI: 15/1 days, Wilcoxon two-sample Test, p=0.0194) and by 4.83 days (95%CI: 10/1 days, Wilcoxon two-sample test, p=0.118) in the case of SARS-CoV-2 as shown in Table 3. The analysis of all naso/oropharyngeal samples collected during prevention phases (during asymptomatic/symptomatic RTI) overall resulted in a difference of - 2.17 ΔCt (95%CI: -4.68/0.34 ΔCt, t-test, p=0.09) in comparison to control (Table S2), matched well with results obtained for acute treatment (>99% virus concentration reduction).

**Table 3.** Time-to-virus clearance (qPCR negative) during treated (EF) and untreated (control) viral symptomatic RTI episodes.

| Time to response (days) | All viral RTI Episodes |  |  | SARS-CoV-2 episodes |  |  |
| --- | --- | --- | --- | --- | --- | --- |
|  | EF | Reference (Control) | p-value<sup>1</sup> | EF | Reference (Control) | p-value<sup>1</sup> |
| n | 8 | 10 |  | 5 | 8 |  |
| Mean | 11.4(2.1) | 19.4(8.6) | 0.019 | 11.8(1.8) | 16.6(6.2) | 0.118 |
| Median | 12 | 18.5 |  | 13 | 16 |  |
| 95%CI for Mean | 9.6/13.2 | 13.3/25.5 |  | 9.6/14.0 | 11.5/21.8 |  |

Data are mean(SD). Analyzable sample sets per day and study groups (n) are indicated. <sup>1</sup>Wilcoxon Two-Sample Test with t-approximation.

### Symptomatic Expression of (viral) symptomatic RTIs and use of co-medication

Compared to control, EF treatment significantly reduced the number of fever days (defined as a temperature of ≥37.8 °C) from 11 (control) to only 1 day in the verum group (RR=0.1, Chi-square test, p=0.0048) and the average body temperature over 6 out of 10 days of acute treatment days significantly (Table S3). Otherwise, no effects on symptom expression were observed. The use of co-medication during RTI episodes was frequent and different in both study groups with 38 incidences of use during 13 RTI episodes in the EF group and 49 incidences during 14 RTI episodes in the control group (ratio: 0.84). It is noteworthy that the use of RTI symptom-related medication (EF: 3, control: 8, ratio: 0.4) was higher in the control group.

### Safety

Overall, 3 and 5 adverse events (AE) were noted for N=3 and N=5 subjects in the EF and control group but none was in relation to study medication and all resolved without sequalae. Notably, out of 5 AEs recorded in the control group, 2 serious COVID-19 illnesses that led to hospitalization as serious adverse events (SAE) were reported but none with *Echinaforc*e, despite the higher rate of co-morbidities in the EF group as shown earlier (Table 1).

## Discussion

The results of this study provide further evidence for antiviral effects of Echinaforce extract (EF) against respiratory viruses, including SARS-CoV2, despite the relatively small sample size and exploratory design.

5 months EF prevention resulted in a 25% infection reduction with any respiratory virus that increased to 43% for enveloped viruses and to 48% for coronaviruses. Interestingly, the strongest risk reduction (63%) was found for infection with SARS-CoV2 viruses, pointing towards a specificity against enveloped viruses overall. The observed protective effect size for SARS-CoV-2 should certainly not be over interpreted, but viewed as further addition in the collation to the significant reduction of SARS-CoV-2 and overall virus loads of more than 2.12log during acute RTI episodes. Although the a priori defined, clinically relevant effect size of 25% was reached at the level of any RTI virus, significance was only attained for the prevention of coronaviruses, and for SARS-CoV-2. Assumptions for the power calculation were based on the pre-pandemic situation and did not take into account containment measures such as disinfection, wearing masks or social distancing. For example, influenza viruses were not observed in the current study and the demonstrated, preventive effects of Echinaforce for this particular virus could not be reproduced [19].

Nevertheless, our results are consistent with, and a further extension of earlier clinical prevention studies comparing Echinaforce extract to control/placebo on endemic RTI viruses. Jawad applied EF extract continuously over 4 months and identified an odds ratio OR =0.49 (p= 0.0114) for infections with enveloped viruses, including endemic coronaviruses such as CoV-229, HKU1 or OC43 [20]. In another study, the same EF extract was administered for 2 × 2 months for prevention in children, interrupted with a one-week treatment break [19]. Consistent with our findings, Ogal (2020) observed significantly fewer infections with enveloped viruses in the Echinaforce group (OR = 0.43, p=0.0038), further substantiating the relevance of antiviral effects *in vivo* [19].

In this study, 1-week breaks succeeded every second prevention month during which no symptomatic RTI episodes occurred but routine testing identified 15 positive PCR/serology tests. In a sensitivity analysis, their consideration for the analysis slightly increased the relative risk with RR=0.68 (95% CI: 0.35/1.32, p>0.05) for SARS-CoV-2 infections. Though not statistically significant, these results might be an indication for quick decline in antiviral effects of *Echinacea* upon treatment cessation. To keep preventive effects high throughout, it may be therefore suggested shortening treatment breaks to a few days, or treating continuously, without treatment breaks. On the rise of the global pandemic, SARS-CoV-2 vaccines have been developed with an extraordinary speed and have mostly proven their effectivity in reducing severe COVID-19 illnesses [21]. Vaccines were also initially found to be effective in reducing peak and overall virus loads more efficiently [22, 23]. A 2.8 – 4.5-fold reduction of peak virus loads in individuals vaccinated against SARS-CoV-2 was reported > 2 weeks post-immunization [23]. This effect apparently reduced over 6 months post-immunization and with increasing activity of the delta-variant [22, 23]. Obviously, there is currently growing interest in additive treatments [24, 25] with proven effectiveness in reducing virus load in the nasal/oral cavity in infected individuals in order to help reduce probability of virus shedding and ultimately transmission [26]. These preparations should ideally be widely available, easy to use and safe[24]. Our findings demonstrate that EF treatment during acute RTI episodes significantly reduced virus loads (all viruses and SARS-CoV-2) by more than 99%. This was further consistent with observations by Nicolussi, (2021) observing a 98.5% reduction on day 2 of illness treated with the same EF preparation (p<0.046) and the shortened time to become virus free (qPCR negative) [14]. Our results represent averages over 5 months of prevention and we did not monitor a potential decay of antiviral effects over time. However, preliminary results suggest that respiratory viruses show limited ability to evade antiviral effects attributed to Echinaforce extract, possibly due to the multicomponent character of plant extractions [12]. This apparently also applies to most relevant SARS-CoV-2 variants of concerns, including the alpha, beta, gamma and delta variants, as a most recent study demonstrated *in vitro* [27]. Potential effects of the EF treatment on the infectiousness of SARS-CoV-2 variants in infected patients are currently being investigated in more detail.

In contrast to the overall symptomatic expression, we observed treatment effects on development of fever and possibly also on severe COVID-19 (hospitalization). The higher rate of concomitant cold medication in the control group could well have masked further effects of the *Echinaforce* treatment on the symptom level requiring confirmation in larger clinical settings.

As mentioned, this study has limitations, first it used descriptive statistical methods, was small in size and secondly it did not use placebo for control and was not blinded. Nevertheless, the design was still considered valid to provide essential evidence for the preventive use of Echinacea during the COVID-19 pandemic for the following reasons: a first parameter was defined as incidence of (viral) RTIs, for which sample size calculation found sufficient statistical power of >80% for 120 included subjects.

The lack of blinding/placebo might be considered a methodological weakness, but it can be assumed that the placebo effect/knowledge of therapy have only limited effects on detection of viral pathogens in NP/OP samples and blood serum. We therefore think that the study design was suitable to address the research question on antiviral effects of Echinaforce *in vivo*.

## Conclusion

A commercial preparation of *Echinacea purpurea* in the licensed dosage (Echinaforce extract), represents a safe, easy-to-use and widely available cost-efficient antiviral with effects in preventing respiratory tract infections, including SARS-CoV2 and reducing virus load. It may add well to existing counter measures in the current COVID-19 pandemic like vaccinations, social distancing and wearing protective facemasks. Future confirmatory studies are warranted.

## Supporting information

Supplementary Results

## Data Availability

All data produced in the present study are available upon reasonable request to the authors.

## Authorship

All named authors meet the ICMJE criteria for authorship for this article, take responsibility for the integrity of the work as a whole, and have given their approval for this version to be published.

## Authors’ contributions

Conceptualization: Giuseppe Gancitano, Lilyana Mircheva, Emil Kolev, Krassimir Kalinov, Wim vanden Berghe; Methodology: Giuseppe Gancitano, Emil Kolev, Lilyana Mircheva, Krassimir Kalinov, Wim vanden Berghe; Formal analysis and investigation: Giuseppe Gancitano, Emil Kolev, Krassimir Kalinov, Lilyana Mircheva, Samo Kreft, Wim vanden Berghe, Michael R. Edwards, Sebastian L. Johnston, Rainer Stange; Writing - original draft preparation: Giuseppe Gancitano, Emil Kolev, Lilyana Mircheva, Krassimir Kalinov, Wim vanden Berghe, Samo Kreft, Michael R. Edwards, Sebastian L. Johnston, Rainer Stange; Writing – review and editing: Giuseppe Gancitano, Emil Kolev, Lilyana Mircheva, Krassimir Kalinov, Wim vanden Berghe, Samo Kreft, Michael R. Edwards, Sebastian L. Johnston, Rainer Stange

## Medical writing, editorial, and other assistance

Support and editorial assistance in the preparation of this article was provided and funded by A. Vogel AG.

## Compliance with ethics guidelines

The Ethics Committee at Diagnostics and Consultation Center Convex Ltd, Sofia, registration nr: 116/26.10.2020) authorized the study protocol (version 1.0, 29. October 2020). Amendments to the study protocol were generated and approved (version 2.0, 17. December 2020, and version 3.0, 14. April 2021). The clinical study is registered at Clinicaltrials.gov (identifier: NCT05002179).

Written informed consent was obtained from all subjects before they participated in the study. Accordance with the ethical principles of the Declaration of Helsinki/Good Clinical Practice guidelines (2013), adherence to national/regional regulatory requirements, and data protection were ensured permanently.

## Data availability

The datasets generated during and/or analyzed during the current study are available from the corresponding author on reasonable request.

