## Supplementary Results for "Echinacea purpurea for the Long-term Prevention of Viral Respiratory Tract Infections during COVID-19 Pandemic: A Randomized, Open, Controlled, Exploratory Clinical Study"

### Authors and institutional addresses

^1^Clinical Research Center DCC Convex Ltd., 11 A Sinanishko ezero, Str. Sofia 1680, Bulgaria

^2^Virtus Respiratory Research Limited, London Bioscience Innovation Centre, 2 Royal College St, London, NW1 0NH, United Kingdom

^3^National Heart Lung Institute, Imperial College London St Marys Campus, Norfolk Place London W2 1PG, United Kingdom.

^4^Medistat Ltd. Statistical Services, Mladost 1A, 1729 Sofia, Bulgaria

^5^Charité – Universitätsmedizin Berlin, Immanuel Hospital Berlin, Königstrasse 63, D-14109 Berlin, Germany

^6^1st  "Tuscania" Paratrooper Regiment Carabinieri, Italian Ministry of Defence, 57127 Livorno, Italy

^7^Laboratory of Protein Chemistry, Proteomics and Epigenetic Signaling (PPES) and Integrated Personalized and Precision Oncology Network (IPPON), Department of Biomedical Sciences, University of Antwerp (UA), Antwerp, Belgium

^8^Faculty of Pharmacy, University of Ljubljana, Aškerčeva cesta 7, Ljubljana, Slovenia

**Clinical Trials registration Nr:** NCT05002179

Supplementary Materials

**Table S1.** Cycle threshold values (Ct) according to qPCR during EF treated (Echinaforce) and untreated (control) viral symptomatic RTI episodes overall and COVID-19 episodes (A). Comparison of virus load reduction between time points per study group (B).

| **(A)** |  | Viruses overall | | | SARS-CoV-2 viruses | | | |
| --- | --- | --- | --- | --- | --- | --- | --- | --- |
|  |  | day 1 | day 5 | day10 | | day 1 | day 5 | day10 |
| EF treatment | n | 9 | 11 | 11 | | 6 | 8 | 8 |
|  | Mean (Ct)^1^ | 21.60(3.07) | 27.82(4.97) | 36.24(7.39) | | 21.34(3.28) | 27.06(3.99) | 32.96(5.73) |
|  | Median (Ct) | 21.4 | 26.79 | 36.19 | | 21.48 | 26 | 32.29 |
|  | 95%CI of Mean | 19.23/23.96 | 24.48/31.16 | 31.28/41.21 | | 17.90/24.78 | 23.72/30.40 | 28.17/37.75 |
| No treatment (control) | n | 11 | 10 | 9 | | 9 | 8 | 7 |
|  | Mean (Ct) ^1^ | 21.52(4.99) | 23.15(4.09) | 29.15(6.31) | | 22.22(4.98) | 23.98(4.17) | 29.75(6.82) |
|  | Median (Ct) | 21.9 | 24.82 | 30.11 | | 22.5 | 25.93 | 30.11 |
|  | 95%CI of Mean | 18.16/24.87 | 20.22/26.07 | 24.30/34.00 | | 18.39/26.05 | 20.5/27.47 | 23.45/36.06 |
| Difference | Mean^2^ (∆Ct) | -0.08 | -4.67 | -7.09 | | 0.88 | -3.08 | -3.21 |
|  | 95%CI of Mean | -3.92/3.76 | -8.82/-0.53 | -13.5/-0.65 | | -3.73/5.49 | -7.45/1.30 | -10.4/3.95 |
|  | p-value^4^ | 0.965 | 0.0292 | 0.0327 | | 0.687 | 0.154 | 0.348 |
| **(B)** |  |  |  |  | |  |  |  |
|  |  | Viruses overall | | | | SARS-CoV-2 viruses | | |
|  |  | day 1 vs. day 5 | day 1 vs day 10 | Day 5 vs day 10 | | day 1 vs. day 5 | day 1 vs day 10 | Day 5 vs day 10 |
| EF treatment | Mean (log∆Ct) ^3^ | -2.19 | -4.73 | -2.54 | | -2.14 | -3.91 | -1.77 |
|  | 95%CI of Mean | -3.08/-1.30 | -6.01/-3.45 | -3.82/-1.25 | | -3.21/-1.06 | -5.14/-2.68 | -2.71/-0.83 |
|  | p-value^2^ | 0.0003 | <0.0001 | 0.0013 | | 0.0022 | 0.0001 | 0.0029 |
| Control | Mean (log∆Ct) ^3^ | -0.07 | -1.91 | -1.84 | | 0.03 | -1.71 | -1.74 |
|  | 95%CI of Mean | -1.04/0.90 | -3.33/-0.49 | -3.58/-0.10 | | -1.10/1.18 | -3.63/0.21 | -4.04/0.56 |
|  | p-value^2^ | 0.8706 | 0.0144 | 0.0403 | | 0.9427 | 0.0724 | 0.1130 |

Data are ^1^mean Ct values(SD) resp. ^2^Mean ∆Ct, resp. ^3^mean log∆Ct values. ^4^Student’s t-test with Satterthwaite modification comparing EF treatment vs control. ^2^Student’s t-test with Satterthwaite modification comparing sampling time points.

**Table S2.** Virus loads (Ct values) during EF treatment/prevention (EF) vs non-treatment (control) as per qPCR.

|  | **all viral detections^2^** | | | **SARS-CoV-2 detections^2^** | | |
| --- | --- | --- | --- | --- | --- | --- |
| **Viral load (Ct):** | EF | Reference (Control) | p-value | EF | Reference (Control) | p-value |
| n | 55 | 84 |  | 31 | 56 |  |
| Mean (Ct) | 30.0(7.6) | 27.8(7.1) | 0.09^4^ | 30.3(7.8) | 29.1(7.5) | 0.459^1^ |
| Median | 29.97 | 27.26 |  | 31.3 | 27.95 |  |
| 95%CI for Mean | 27.94/32.07 | 26.29/29.38 |  | 27.44/33.17 | 27.05/31.05 |  |

Data are mean(SD). Analyzable sample sets per day and study groups (n) are indicated. ^1^Student’s t-test

**Table S3**. Course of body temperatures (°C)/fever days (>37.8 °C) during EF treated (Echinaforce) and untreated (control) viral symptomatic RTI episodes overall

| EF treatment | day | 1 | 2 | 3 | 4 | 5 | 6 | 7 | 8 | 9 | 10 |
| --- | --- | --- | --- | --- | --- | --- | --- | --- | --- | --- | --- |
|  | Mean (°C) | 36.4 (0.5) | 36.4 (0.3) | 36.6 (0.5) | 36.4 (0.4) | 36.4 (0.5) | 36.3 (0.4) | 36.2 (0.5) | 36.2 (0.4) | 36.3 (0.4) | 36.3 (0.5) |
|  | Median | 36.5 | 36.4 | 36.4 | 36.5 | 36.4 | 36.4 | 36.5 | 36.4 | 36.4 | 36.5 |
|  | Fever days (N)^2^ | 0 | 0 | 1 | 0 | 0 | 0 | 0 | 0 | 0 | 0 |
| No treatment (control) | day | 1 | 2 | 3 | 4 | 5 | 6 | 7 | 8 | 9 | 10 |
|  | Mean (°C) | 37.4(0.9) | 37.2(0.7) | 36.9(1.0) | 36.7(0.8) | 36.8(0.6) | 36.7(0.4) | 36.6(0.5) | 36.6(0.3) | 36.5(0.4) | 36.5(0.3) |
|  | Median | 37.3 | 37.2 | 36.6 | 36.5 | 36.6 | 36.6 | 36.5 | 36.7 | 36.5 | 36.4 |
|  | Fever days (N) ^2^ | 4 | 3 | 1 | 1 | 2 | 0 | 0 | 0 | 0 | 0 |
|  | p-value^1^ | 0.002 | 0.003 | 0.326 | 0.128 | 0.039 | 0.033 | 0.034 | 0.013 | 0.22 | 0.174 |

Data are mean(SD). ^1^Student’s t-test with Satterthwaite modification, ^2^Data are N=fever days (≥37.8 °C).
